# Variation in the reporting of elective surgeries and its influence on patient safety indicators

**DOI:** 10.1101/2021.05.29.21257635

**Authors:** Kenneth J. Locey, Thomas A. Webb, Sana Farooqui, Bala Hota

## Abstract

**Background:** US hospital safety is routinely measured via patient safety indicators (PSIs). Receiving a score for most PSIs requires a minimum number of qualifying cases, which are partly determined by whether the associated diagnosis-related group (DRG) was surgical and whether the surgery was elective. While these criteria can exempt hospitals from PSIs, it remains to be seen whether exemption is driven by low volume, small numbers of DRGs, or perhaps, policies that determine how procedures are classified as elective.

**Methods:** Using Medicare inpatient claims data from 4,069 hospitals between 2015 and 2017, we examined how percentages of elective procedures relate to numbers of surgical claims and surgical DRGs. We used a combination of quantile regression and machine learning based anomaly detection to characterize these relationships and identify outliers. We then used a set of machine learning algorithms to test whether outliers were explained by the DRGs they reported.

**Results:** Average percentages of elective procedures generally decreased from 100% to 60% in relation to the number of surgical claims and the number of DRGs among them. Some providers with high volumes of claims had anomalously low percentages of elective procedures (5% – 40%). These low elective outliers were not explained by the particular surgical DRGs among their claims. However, among hospitals exempted from PSIs, those with the greatest volume of claims were always low elective outliers.

**Conclusion:** Some hospitals with relatively high numbers of surgical claims may have classified procedures as non-elective in a way that ultimately exempted them from certain PSIs.

## Background

From individual practices to global networks, modern healthcare has been transformed by the need to generate, accumulate, and analyze data.^1-3^ In addition to shaping the quality and safety of practice, and the policies that govern it, this data revolution has created new dimensions of opportunity.^4^ Healthcare providers and researchers can now ask and answer questions that were previously untenable by plying powerful analyses to immense data sets.^5^ Within studies of quality and patient safety, analyses can now go beyond the examination of scores and benchmarks by digging deeper into sources of variation among thousands of providers across multiple years. To complement this endeavor, studies have the potential to leverage the increasing availability of data science tools that are purpose-built to find telling patterns within large and complex data sets. However, studies of quality and patient safety that are granular, longitudinal, national in scale, and that employ advanced analytics (e.g., machine learning) are still relatively uncommon.

Across the US, hospital safety is routinely measured via patient safety indicators (PSIs), 26 metrics developed by the Agency for Healthcare Research and Quality (AHRQ).^6^ Calculations of most PSIs are based on medical claims submitted for various diagnosis-related groups (DRGs), an inpatient classification system established by The Centers for Medicare and Medicaid Services (CMS).^7^ Put simply, the number of claims for qualifying DRGs form the denominators of PSIs and the number of adverse safety events coded among those claims form PSI numerators.^8^ The inclusion of a claim into a PSI can depend on whether the associated DRG is classified as surgical and, if so, whether the surgery was classified as elective or non-elective (Table 1). PSI scores vary greatly among hospitals and may be driven by the patient population and volume, as well as hospital accreditation and safety culture.^9-11^ However, underlying the factors leading to variation in PSI scores are the factors influencing variation in PSI denominators and whether a hospital receives a score at all.

**Table 1.**
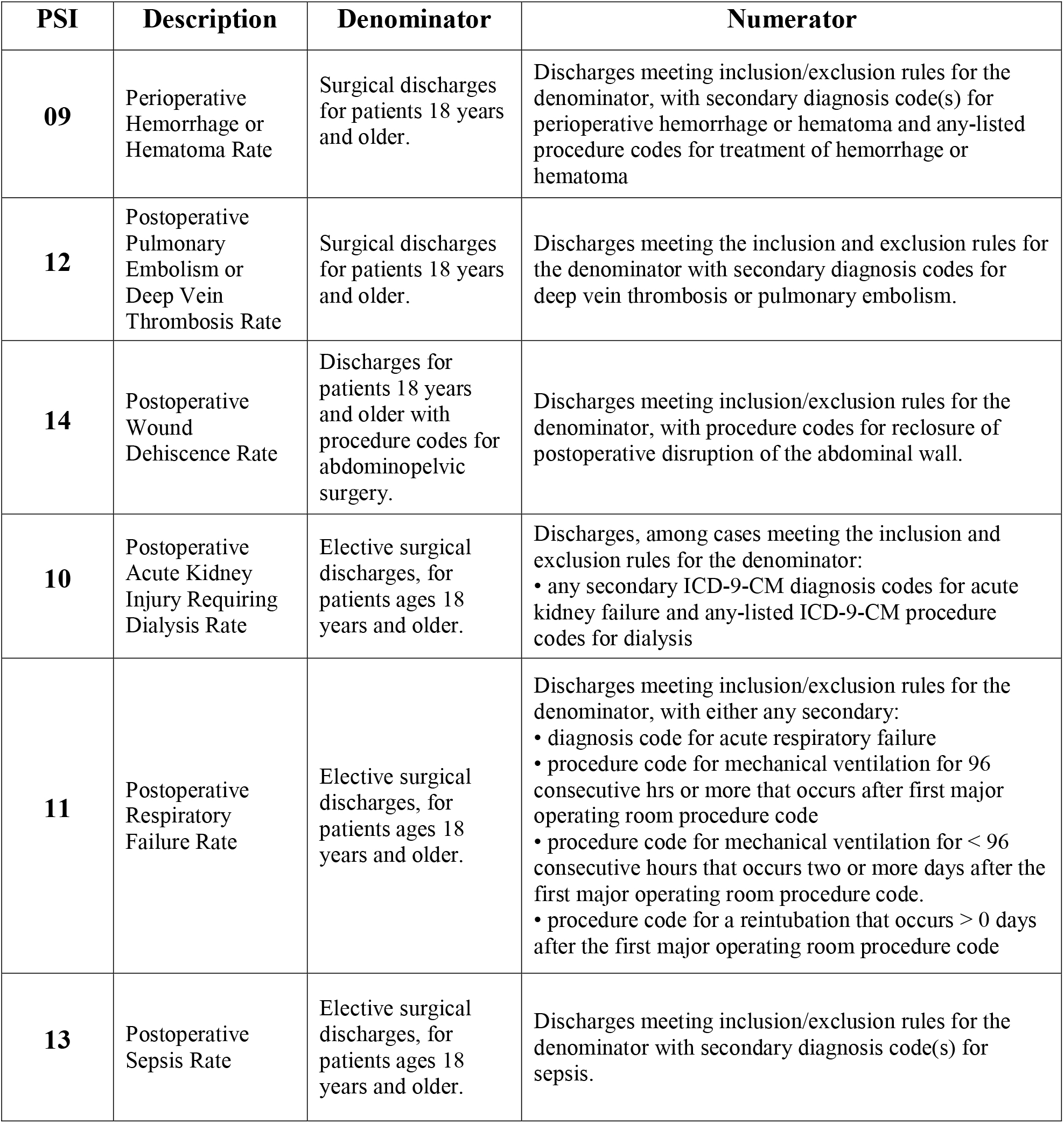
Descriptions of patient safety indicators (PSI) in the present work. The AHRQ website offers greater details on numerators and denominators (https://www.qualityindicators.ahrq.gov).

| PSI | Description | Denominator | Numerator |
| --- | --- | --- | --- |
| **09** | Perioperative Hemorrhage or Hematoma Rate | Surgical discharges for patients 18 years and older. | Discharges meeting inclusion/exclusion rules for the denominator, with secondary diagnosis code(s) for perioperative hemorrhage or hematoma and any-listed procedure codes for treatment of hemorrhage or hematoma |
| **12** | Postoperative Pulmonary Embolism or Deep Vein Thrombosis Rate | Surgical discharges for patients 18 years and older. | Discharges meeting the inclusion and exclusion rules for the denominator with secondary diagnosis codes for deep vein thrombosis or pulmonary embolism. |
| **14** | Postoperative Wound Dehiscence Rate | Discharges for patients 18 years and older with procedure codes for abdominopelvic surgery. | Discharges meeting inclusion/exclusion rules for the denominator, with procedure codes for reclosure of postoperative disruption of the abdominal wall. |
| **10** | Postoperative Acute Kidney Injury Requiring Dialysis Rate | Elective surgical discharges, for patients ages 18 years and older. | Discharges, among cases meeting the inclusion and exclusion rules for the denominator:<br>• any secondary ICD-9-CM diagnosis codes for acute kidney failure and any-listed ICD-9-CM procedure codes for dialysis |
| **11** | Postoperative Respiratory Failure Rate | Elective surgical discharges, for patients ages 18 years and older. | Discharges meeting inclusion/exclusion rules for the denominator, with either any secondary:<br>• diagnosis code for acute respiratory failure<br>• procedure code for mechanical ventilation for 96 consecutive hrs or more that occurs after first major operating room procedure code<br>• procedure code for mechanical ventilation for < 96 consecutive hours that occurs two or more days after the first major operating room procedure code.<br>• procedure code for a reintubation that occurs > 0 days after the first major operating room procedure code |
| **13** | Postoperative Sepsis Rate | Elective surgical discharges, for patients ages 18 years and older. | Discharges meeting inclusion/exclusion rules for the denominator with secondary diagnosis code(s) for sepsis. |

Publicly available PSI data reveal that hospitals with PSI denominators less than 25 are excluded from certain PSI calculations.^12^ This minimum cutoff is intended to prevent random effects of small sample sizes on the frequency of adverse safety events.^13^ Though reasonable from a statistical perspective, this rule allows the classification of procedures as elective or non-elective to influence the size of PSI denominators and, hence, whether hospitals are exempted from a particular PSI score. Specifically, when considering the inclusion rules for PSI denominators, classifying surgical procedures as elective can never decrease PSI denominators but classifying them as non-elective can (Table 1). PSI-related concerns over the effects of classifying procedures as elective or non-elective have been raised by others.^14^ However, it remains to be seen whether the tendency to report low denominators for elective-based PSIs, or to be excluded from them, is simply driven by the volume of surgical claims and the various surgical DRGs among them.

In the present study, we use three years of CMS claims data from 4,069 hospitals to examine trends in the percent of elective procedures performed across surgical DRGs. We characterize how the percent of elective surgeries relates to the total numbers of surgical claims and the number of surgical DRGs for which hospitals submit claims. We then use machine learning-based dimensionality reduction and clustering to determine whether the specific surgical DRGs for which hospitals submit claims explain why some hospitals have consistently low percentages of elective surgeries. Finally, we ask whether or not hospitals with reasonably high volumes of surgical claims have consistently been exempted from elective based PSIs by reporting anomalously low percentages of elective surgeries. We discuss the implications of our findings on PSI outcomes, uncertainties in the reporting of elective surgeries, and the benefits of combining large granular data sets and advanced data science methods for identifying general trends and outliers in the area of quality and safety analytics.

## Methods

### Data

We used three consecutive years (2015 – 2017) of Medicare inpatient claims data from CMS’s Limited Data Set (LDS) files containing individual claims data from 5,486 hospitals. We accessed these data through services provided by CareJourney (https://carejourney.com/), a healthcare analytics company, and our longitudinal span was determined by the data available to us. Within the LDS data, claims are indicated as surgical (P) or medical (M) and as either elective (TYPE_ADM = 3), emergency, urgent, newborn, trauma center, or unknown.^15^ We extracted hospital-specific variables including total annual claims for surgical DRGs, annual number of surgical DRGs for which claims were submitted, annual number of claims per surgical DRG, and the annual percent of claims reported as elective procedures for each surgical DRG. In compliance with user agreements, we excluded all hospital-level DRG records containing either less than 11 claims or where the percent of elective procedures for a given DRG could be used to back calculate a number of claims less than 11. In accordance with CMS and CareJourney data use agreements, we cannot provide raw data but, upon request, will provide permissible secondary data with de-identified providers and all source code needed to reproduce our analyses and figures.

We obtained patient safety indicator (PSI) data spanning measurement dates between 2014 and 2018 from archived Complications and Deaths files, now available at CMS Hospital Compare (https://data.cms.gov/provider-data/archived-data/hospitals); also obtained in March 2020. We avoided naming providers by replacing provider CMS numbers with randomly generated values and by removing names, states, addresses, and all other provider-level information that was not necessary to our analyses. However, because our analyses intersect LDS data with Hospital Compare data, identification is theoretically possible.

### Analyses

#### Average percent of elective surgeries (APE)

We calculated the average percent of claims reported as elective procedures across DRGs (APE) for each provider, for each year:

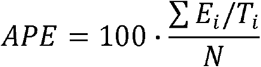

Where *E*_*i*_ is the number of claims for the *i*^*th*^ DRG that were reported as elective procedures, *T*_*i*_ is the total number of claims reported for the *i*^*th*^ DRG, and *N* is the number of DRGs reported. This calculation gives equal weight to each DRG reported by a hospital. In contrast, dividing the total number of elective procedures by the total number of claims would give greater weight to common DRGs and be less sensitive to variations among less common DRGs.

#### APE vs. number of claims and DRGs

We examined relationships between APE and 1) the total number of surgical DRG claims (i.e., total claims) reported by each provider in each year (2015 – 2017) and 2) the number of surgical DRGs reported by each provider in each year. We used polynomial quantile regression to characterize these relationships and to construct bands outside of which potential outliers would be apparent. This approach allows quantile regression to capture curvilinear trends across entire datasets and across quantiles of the data distribution.^16^ For example, the 0.02 quantile (analogous to the 2^nd^ percentile) may capture a curvilinear trend among data points located near the bottom of a relationship while the 0.98 quantile (98^th^ percentile) may capture a different curvilinear trend near the top of a relationship. Studies often use percentiles to refer to the results of quantile regression.^16^ We conducted these analyses using the python-based statsmodels library.

#### Outlier detection

We classified providers who demonstrated consistently low or consistently high anomalous APE values as ‘APE outliers’. We did this using a cautious set of criteria based on all three years of provider data and with respect to the relationship of APE to both the number of annual claims for surgical DRGs and the number of surgical DRGs among those claims. Specifically, we used a combination of polynomial quantiles and formal anomaly detection via isolation forest.^17^ Isolation forest is a formal machine learning-based outlier detection technique based on the concept of random forests, a commonly used machine learning-based classification technique.^17^ We conducted isolation forest analyses, and all other machine learning analyses using the python-based machine learning library scikit-learn.

Using polynomial quantile regression, we considered providers as low APE outliers if their APE fell below the 2^nd^ percentile (0.02 quantile) in each relationship (APE vs. total claims, APE vs. no. of DRGs), for each year. This forgiving set of criteria meant that each provider only had to fall above the 2^nd^ percentile for one year in any of the two relationships, to not be considered a low APE outlier. High APE outliers were identified in a similar fashion but were based on whether they consistently fell above the 98^th^ percentile (0.98 quantile).

Using isolation forest anomaly detection, we identified providers as outliers if they were classified as such across all years of their data and in both relationships (APE vs. no. of claims, APE vs. no. of DRGs). Again, these criteria meant that each provider only had to be classified as a non-outlier for one year in any of the two relationships, to not be considered an APE outlier. Combining results from polynomial quantile regression and isolated forest anomaly detection, providers who were identified as outliers, were classified as such by each of two methods applied to each of two relationships, across all years for which their data was available.

The conservative nature of our approach was three-fold. First, the use of two independent methods, two relationships, and multiple years of data represents a cautious methodology. Second, our approach ensured that providers were not considered outliers for having a single ‘fluke’ year in regards to a single relationship. Third, while our approach identified a smaller number of outliers than might otherwise be found with a less cautious approach, it ensured that providers who were ultimately identified as APE outliers were consistent outliers.

#### Analysis of DRG profiles

We considered that APE outliers may tend to report claims for DRGs for which a low percent of elective surgeries is common. To this end, we asked whether APE outliers can be explained by their DRG profiles, i.e., the set of surgical DRGs for which providers submit claims. Comparing thousands of providers based on hundreds of DRGs involves two common tasks of machine learning: 1) Dimensionality reduction, i.e., reducing a data set with many variables to a few composite dimensions that retain the majority of variation in the original data, and 2) Classification, or clustering data into groups. These tasks are often combined. For example, dimensionality reduction allows clustering algorithms to operate on a smaller number of variables that capture the majority of variation among samples and to ignore variables that are redundant or that simply add noise.^18^ Alternatively, when data have no obvious coordinates for visualization, dimensionality reduction can allow clusters to be visualized in two-dimensional space.^19^

We used principal component analysis (PCA) to reduce the number of explanatory variables from 377 DRGs to the 10 principal components that best explain the variation among provider DRG profiles. PCA is a long-established unsupervised dimensionality reduction technique that uses eigenanalysis (mathematical operations on a square matrix) to generate composite variables (principal components) from linear combinations of the original variables.^20-21^ By finding principal component axes that explain as much variation in the original data as possible, PCA attempts to preserve the explainable variation of the original data and the overall (i.e., global) data structure. This last aspect is considered crucial when clustering on the outputs of dimensionality reduction, making PCA a popular choice for this task.^18,21^

We performed clustering on the first 10 components of PCA using density-based spatial clustering of applications with noise (DBSCAN).^22^ DBSCAN is a machine learning algorithm that attempts to find areas within data that satisfy a minimum density of points (i.e., clusters), which are separated from other clusters by areas of lower density.^22,23^ DBSCAN identifies core points within data, and all points within a prescribed radius of a core point are considered to be part of the same cluster. Points which are not “density reachable” from any core point are not classified into any cluster.^22,23^ Unlike other clustering methods, DBSCAN is scalable to large samples, does not require the number of clusters as an input, can detect clusters of any shape, and will not force each point into a cluster.

Though DBSCAN does not require the number of clusters to be predetermined, it is not without parameters. These are: 1) ε, the maximum distance between two points for one to be considered as being in the neighborhood of the other, and 2) MinPts, the minimum number of points in a neighborhood needed to determine a core point. While general heuristics are available for choosing values for ε and MinPts ^24^, we explored a range of parameter value combinations to detect whether or not combinations of ε and MinPts lead DBSCAN to generate clusters that support that APE outliers can be explained by their DRG profiles. Since components of PCA are orthogonal (mathematically independent of each other), we used Euclidean distance as our DBSCAN distance metric.

Because DRG profile data have no inherent x-y coordinates by which to visualize clusters (as in spatial or image data) we visualized the resulting clusters of DBSCAN by performing dimensionality reduction on provider DRG profiles using a technique that is specifically designed for visualization, i.e., t-distributed stochastic neighbor embedding (t-SNE).^25^ In short, t-SNE converts similarities between data points to joint probabilities and attempts to minimize the relative entropy (i.e., how different one probability distribution is from a second) between the joint probabilities of low-dimensional embedding (e.g., 2-dimensional depiction) and the high-dimensional data (i.e., 377 DRGs among several thousand providers). Whereas PCA is a deterministic linear method that preserves global data structure, t-SNE is a nonlinear probabilistic method that preserves local data structure (positions of points within clusters).These attributes of t-SNE allow it to produce visualizations with high separation between clusters and high fidelity of samples within clusters. DBSCAN and t-SNE are often used in combination, i.e., DBSCAN to assign data to clusters and t-SNE to visualize data in low-dimensional space.^26-27^

#### PSI exemption

PSIs based on surgical procedures, elective or otherwise, are not reported when the qualifying volume of claims is less than 25.^12^ Consequently, low-volume hospitals stand a greater chance of being exempted from PSI reporting. However, hospitals with a reasonably high volume of surgery-related claims that report exceptionally low percentages of elective surgeries may also stand a greater chance of being excluded from PSI reporting. To test this, we examined the tendency for hospitals to be excluded from six PSIs based on surgical procedures (Table 1). Three of these PSI’s are inclusive of elective and non-elective procedures (PSIs 09, 12, 14) and three only include elective procedures (PSIs 10, 11, 13) (Table 1). Using the footnotes column associated with PSI data, we quantified how many PSIs were excluded for each hospital based on the following footnotes: “1 - The number of cases/patients is too few to report”, “7 - No cases met the criteria for this measure.”

Because PSI measurements span multiple consecutive years, we used PSI measurement dates having midpoint years corresponding to the years in our claims data (e.g., 2015 - 2017). For example, if the measurement time span ranged from 1 April 2015 to 31 March 2018, then using the median date (29 September 2016), we inferred the midpoint measurement year to be 2016. This approach cannot produce a perfect corollary but, by using the midpoint year, may be more accurate than using the start year or end year.

## Data and source code availability

Upon request, we will provide all permissible data and all source code for reproducing our analyses and figures. Our source code was written in the Python programming language (v3.7) and uses common python libraries (e.g., numpy, pandas, scipy) and the python-based scikit-learn machine learning library.

## Results

### APE vs. number of claims and DRGs

Among providers, the average of the percent of claims reported as elective procedures across DRGs (APE) generally decreased from ∼100% to ∼60% in relation to the total number of surgical claims and the number of surgical DRGs that providers reported (Fig 1). In general, a greater total number of surgical claims and a greater number of DRGs among those claims both led to a lower APE (Fig 1). The vast majority of providers followed these general trends and maintained an APE greater than 50%. However, the variation in these relationships was also substantial, as revealed by the varied forms of 98^th^ and 2^nd^ percentiles of a 3^rd^ degree polynomial quantile regression (Fig 1). In particular, a small fraction of providers consistently fell below the 2^nd^ percentile of each relationship (Fig 1). Additionally, while providers that reported claims for relatively low numbers of surgical DRGs tended to have high APE values often approaching 100%, a small fraction of providers with similarly low numbers of surgical DRGs tended towards the opposite direction and had consistently and exceptionally low APE values (Fig 1).

**Figure 1.**
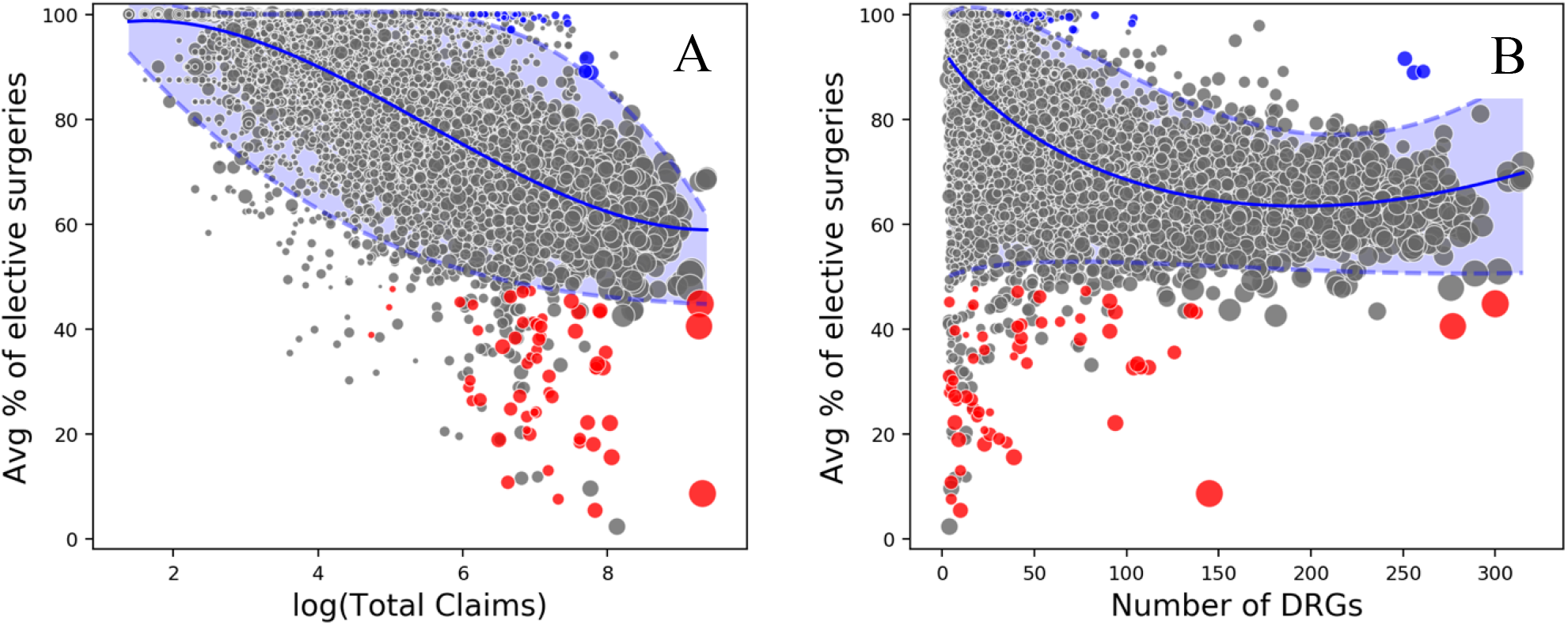
**A**) The relationship between the average of the percent of electives surgeries reported among DRGs (APE) to the total number of claims reported by providers that reported claims for at least four DRGs in each year, 2015 to 2017 (*n* = 4,069 providers). Total numbers of claims are log-transformed. **B**) The relationship between APE and the number of DRGs reported among the 4,069 providers in A. For both A and B, solid blue lines are the fit of polynomial quantile regression to the quantile of least absolute deviation (LAD). Lower and upper dashed blue lines are the 2^nd^ and 98^th^ quantiles, respectively. Red and blue circles are providers that were classified as consistently low or consistently high APE outliers, respectively, via the combination of polynomial quantile regression and isolated forest anomaly detection conducted across all three years of provider data and both relationships (i.e., A & B).

### APE outliers

The combination of polynomial quantile regression, isolated forest anomaly detection, and a set of 12 criteria necessary to be classified as an outlier provider revealed 20 providers that consistently had anomalously low APE values and 9 providers with consistently high APE values (Fig 1). Due to our conservative methodology, an attempt to err on the side of detecting true positives, not meeting all 12 criteria would have caused some providers that may be actual outliers to not be classified as such. Both high and low APE outliers tended to submit a relatively large number of claims (Fig 1), causing us to doubt any potential influence of random effects due to small sample sizes, especially in regards to low APE outliers. Though high and low APE outliers often submitted claims for a relatively small number of DRGs, this was the case for providers, in general.

### Analysis of DRG profiles

The combination of principle component analysis (PCA), density-based spatial clustering of applications with noise (DBSCAN), and t-distributed stochastic neighbor embedding (t-SNE) revealed no tendency for low APE outliers to group together by their DRGs and revealed that most outliers were as similar in their DRGs profiles to non-outliers as they were to other outliers (Fig 2). The visualization of dimension reduction largely reflected the outcome of clustering, an agreement that reinforced our general finding that the DRGs reported by a particular provider did not influence their status as an outlier in the reporting of elective surgical procedures.

**Figure 2.**
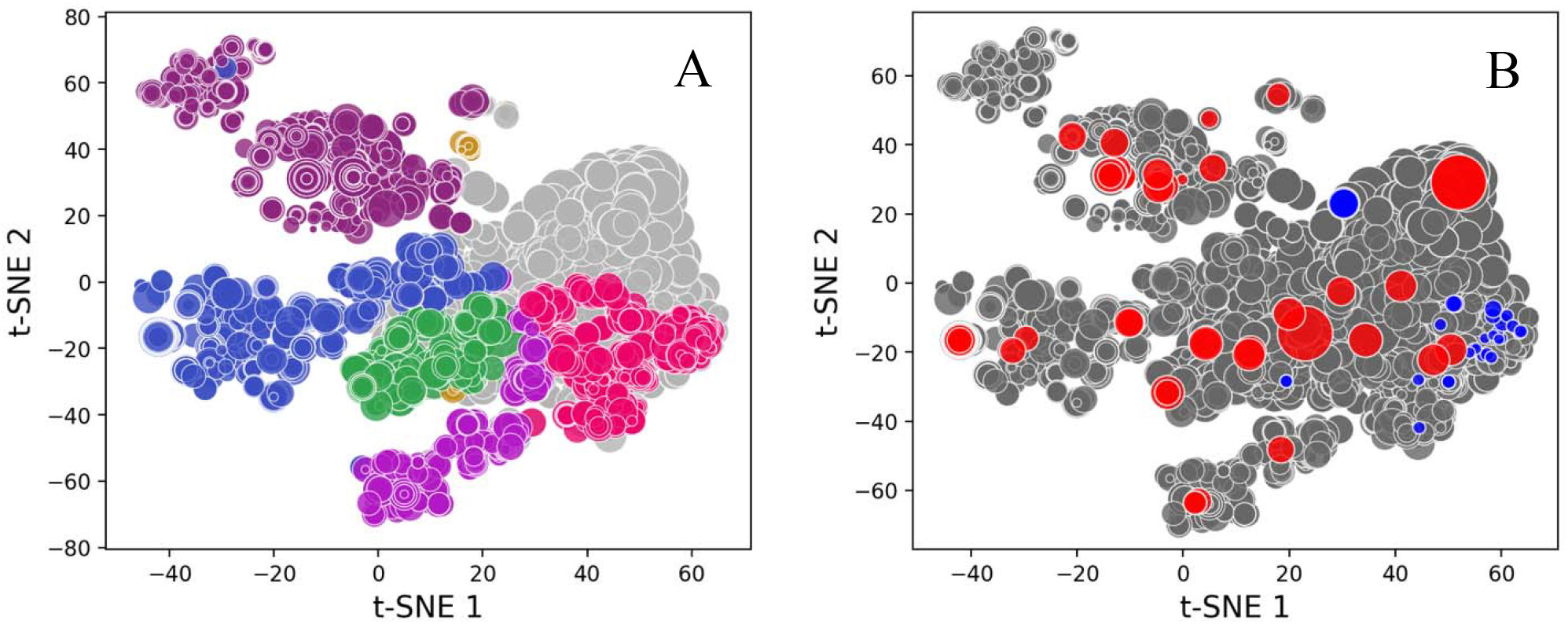
**A**) Combined visualization of dimension reduction and clustering. Each provider that reported claims for at least four DRGs in the years 2015 – 2017 (*n* = 4,069) is represented by a circle with the color corresponding to the cluster assigned by density-based spatial clustering of applications with noise (DBSCAN). Gray circles represent providers that were not assigned to a cluster. Similarities among providers within clusters are represented by the axes of dimension reduction via t-distributed stochastic neighbor embedding (t-SNE). **B**) Visualization via t-SNE, as in A, but with low and high APE outliers are colored by red and blue, respectively. Gray circles are providers that were not classified as APE outliers.

### PSI exemption

As expected, hospitals with relatively low volumes of surgical claims qualifying for PSI inclusion were more likely than others to be excluded from one or more PSI calculations (Fig 3). Hospitals with consistent and anomalously low average percentages of elective surgical procedures (low APE outliers) were rarely excluded from PSI calculations based on all qualifying surgeries, elective or otherwise (Fig 3a). However, these low APE outliers were frequently excluded from one or more PSIs based on elective surgeries, despite their relatively high volumes of surgical claims (Fig 3b). In contrast, and as shown in Figure 1, while hospitals with relatively low volumes of surgical claims also tend to be exempted from PSI calculations, they also tend to report high percentages of elective surgical procedures.

**Figure 3.**
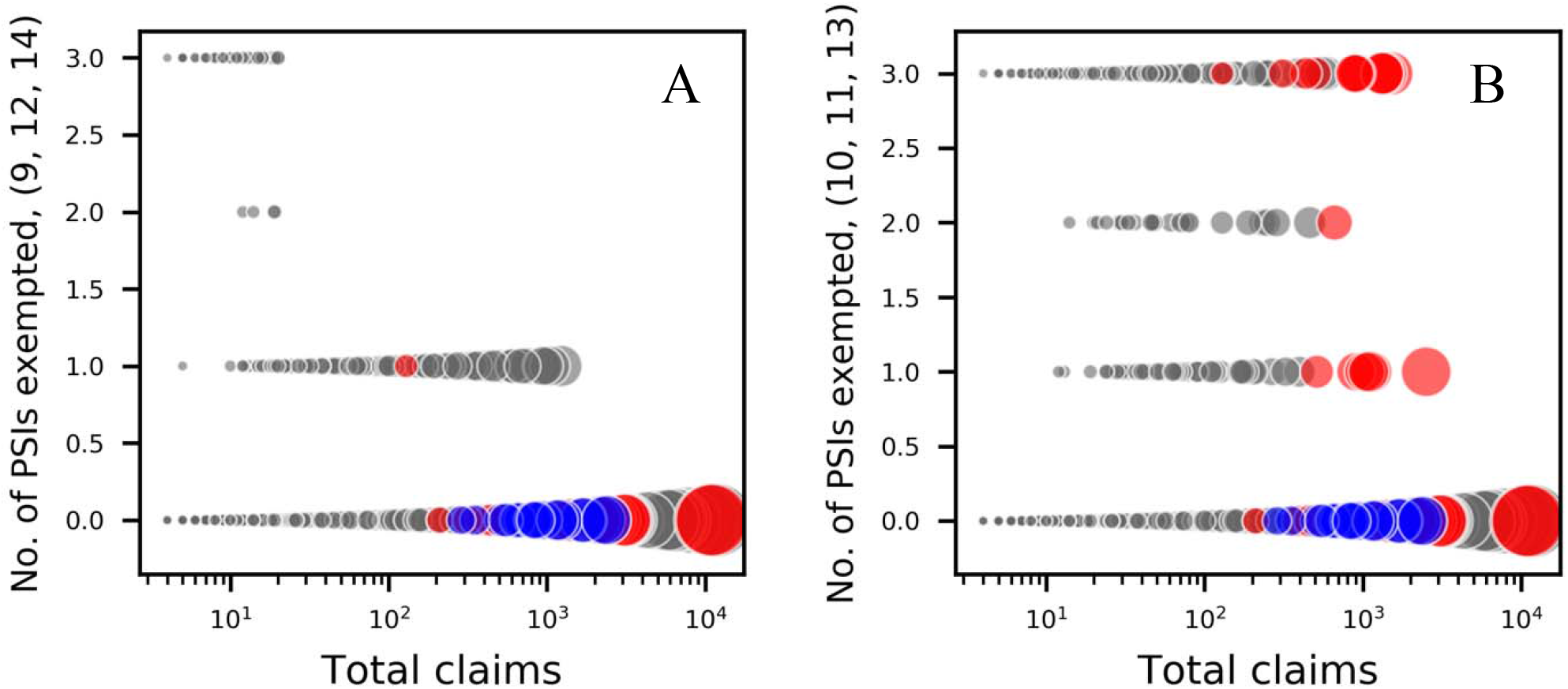
PSI exemptions vs. the total volume of surgical procedures. PSI data are based on midpoint measurement years that correspond to years within CMS LDS claims data (2015 – 2017). Of the 4,069 providers in LDS data having reported at least four DRGs in each year, 578 providers had no corresponding records in PSI data, resulting in a total sample size of 3,491 providers. **A)** Results for PSIs calculated from qualifying cases of elective and non-elective surgical procedures. **B)** Results for PSIs calculated only from qualifying cases of elective surgical procedures. Low and high APE outliers are colored by red and blue, respectively. Gray circles are providers that were not classified as APE outliers.

## Discussion

The reporting of healthcare claims data has profound financial impacts on hospitals and underpins the quality and safety measures influencing hospital rankings and public perception. In similar fashion, the volume of elective surgeries performed within a hospital has a direct impact on the calculation of patient safety indicators (PSIs). In the current study, we examined variation among healthcare providers in regards to the average percent of surgery-related Medicare claims they report as elective procedures (APE). We examined how APE relates to the total number of surgical claims reported and the number and identities of surgical DRGs among those claims. After analyzing PSI and claims data from several thousand providers across three years (2015 – 2017), we uncovered consistent outliers that defy trends in how APE relates to the volume of surgical claims and the number and diversity of surgical DRGs used in PSI calculations. Despite their considerable volumes of claims, these outliers tend to be exempt from reporting of PSIs based on elective surgeries.

To explain why some providers have exceptionally low or high percentages of elective procedures across their surgical DRGs, we considered that some surgical DRGs tend to be reported as elective more frequently than others. Unlike variables such as hospital type, geography, and demography, DRGs directly reflect the procedures performed and billed by hospitals. If hospitals of similar type (e.g., cancer hospitals, pediatric centers, community hospitals) perform generally similar procedures, then our DRG-based analysis would have captured this. But, despite the combination of machine learning designed to identify similarities and natural groupings within complex data (hundreds DRGs among thousands of providers), low APE outliers exhibited no particular similarities in the DRGs for which they submitted claims. Despite the general trends we uncovered, neither the volume of procedures, diversity of procedures, nor the specific sets of DRGs reported by providers gave any insight into the factors contributing to an anomalously low percentages of elective procedures reported by some hospitals. Consequently, sources of variation in the reporting of elective procedures may be found in the reporting and coding practices of individual providers.

Our results suggest that some hospitals with a reasonably large number of surgical claims may have classified surgical procedures as non-elective in a way that differs from other hospitals. However, our results should not be interpreted to imply that some providers are attempting to avoid certain PSI calculations by using classification rules that largely differ from others. Differences in classification rules have been discussed by others and related to differences in the percent of elective surgeries that ultimately influence PSIs.^14^ But, it may also be the case that the percent of elective procedures reported by some providers actually is exceptionally low across their surgical DRGs, despite a relatively high volume of claims. Though we found no evidence to support this alternative explanation, volumes of elective surgeries are known to be sensitive to economic downturn and public health crises.^28,29^ Still, among the hospitals that were exempted from PSI reporting based on elective surgeries, those with the greatest volume of claims were nearly always those that reported anomalously low percentages of elective surgeries.

Throughout our study, we aimed to demonstrate how combinations of machine learning and uncommon regression models can be used in the study of quality and patient safety, particularly when faced with immense multivariate data sets. However, beyond the need to match “big data” with appropriately powerful tools, the study of quality and patient safety is also challenged by the unintended effects of how quality and safety measures are calculated. For example, while a minimum cutoff for PSI denominator values may prevent random effects of small sample sizes, no such cutoff exists for other quality measures, e.g., those related to hospital associated infections (HAIs). Consequently, while the presence of a minimum cutoff can allow hospitals (even those of reasonably high volume) to avoid PSIs, the absence of a minimum cutoff for HAIs should, by the same logic used to calculate PSIs, cause hospitals with low volume to be particularly vulnerable to random effects.

In addition to shedding greater light on the sources of variation among healthcare providers and the potential shortcomings of quality and safety methodologies, we expect that future studies will also overcome other limitations of our research. Specifically, studies will be aided by greater access to additional years of CMS LDS data and the eventual integration of large and granular data sets from different sources. For example, the Healthcare Cost Report Information System (HCRIS) data set maintained by CMS contains several thousand cost and facility related variables for thousands of hospitals, and for each year since 1996 (see: https://www.cms.gov/Research-Statistics-Data-and-Systems/Downloadable-Public-Use-Files/Cost-Reports). The size, complexity, and proprietary file formats of HCRIS data can be prohibitively challenging for potential users, resulting in cottage industry of third-party providers. However, the integration of HCRIS data with LDS data sets as well as quality and safety data from AHRQ would represent a formidable data ensemble, ripe for the application of cutting-edge data science. However, without greater knowledge of the varied policies used by providers to code and classify procedures, coupling powerful tools with immense data may still yield limited insights.

## Data Availability

We will provide all permissible data and source code needed to reproduce our analyses and figures upon request.

## Acknowledgments

We thank the healthcare data analytics service CareJourney for their timely provision of CMS LDS files and assistance.

## Notes

### Competing Interest Statement

The authors have declared no competing interest.

### Clinical Trial

Our study did not involve clinical trials.

### Funding Statement

No external funding was received for this work.

### Author Declarations

Approval to use LDS data from the Centers for Medicare and Medicaid Services (CMS) in the present work was granted by CMS under a Data Use Agreement (LDSS-2019-52808) to NavHealth (CareJourney) who, under their corresponding research proposal, are permitted to share aggregated LDS with our hospital.

## References

1. Murdoch, T.B. and Detsky, A.S., 2013. The inevitable application of big data to health care. Jama, 309(13), pp. 1351–1352.

2. Obermeyer, Z. and Emanuel, E.J., 2016. Predicting the future—big data, machine learning, and clinical medicine. The New England journal of medicine, 375(13), p. 1216.

3. Ilomäki, J., Bell, J.S., Chan, A.Y., Tolppanen, A.M., Luo, H., Wei, L., Lai, E.C.C., Shin, J.Y., De Paoli, G., Pajouheshnia, R. and Ho, F.K., 2020. Application of Healthcare ‘Big Data’in CNS Drug Research: The Example of the Neurological and mental health Global Epidemiology Network (NeuroGEN). CNS drugs, 34(9), pp. 897–913.

4. Raghupathi, W. and Raghupathi, V., 2014. Big data analytics in healthcare: promise and potential. Health information science and systems, 2(1), pp. 1–10.

5. Bragazzi, N.L., Dai, H., Damiani, G., Behzadifar, M., Martini, M. and Wu, J., 2020. How big data and artificial intelligence can help better manage the COVID-19 pandemic. International journal of environmental research and public health, 17(9), p. 3176.

6. Agency for Healthcare Research and Quality. 2015. AHRQ Quality Indicators: Patient Safety Indicators. https://www.qualityindicators.ahrq.gov/Downloads/Modules/PSI/V50/PSI_Brochure.pdf

7. Centers for Medicare and Medicaid Services. 2019. Design and development of the Diagnosis Related Group (DRG). PBL-038. https://www.cms.gov/icd10m/version37-fullcode-cms/fullcode_cms/Design_and_development_of_the_Diagnosis_Related_Group_(DRGs).pdf

8. Agency for Healthcare Research and Quality. 2017. Patient Safety Indicator 09 (PSI 09) Perioperative Hemorrhage or Hematoma Rate. https://www.qualityindicators.ahrq.gov/Downloads/Modules/PSI/V60-ICD09/TechSpecs/PSI_09_Perioperative_Hemorrhage_or_Hematoma_Rate.pdf

9. Mardon, R.E., Khanna, K., Sorra, J., Dyer, N. and Famolaro, T., 2010. Exploring relationships between hospital patient safety culture and adverse events. Journal of patient safety, 6(4), pp. 226–232.

10. Rajaram, R., Chung, J.W., Kinnier, C.V., Barnard, C., Mohanty, S., Pavey, E.S., McHugh, M.C. and Bilimoria, K.Y., 2015. Hospital characteristics associated with penalties in the centers for medicare & medicaid services hospital-acquired condition reduction program. Jama, 314(4), pp. 375–383.

11. Scali, S.T., Giles, K.A., Kubilis, P., Beck, A.W., Crippen, C.J., Hughes, S.J., Huber, T.S., Upchurch Jr, G.R. and Stone, D.H., 2020. Impact of hospital volume on patient safety indicators and failure to rescue following open aortic aneurysm repair. Journal of vascular surgery, 71(4), pp. 1135–1146.

12. Centers for Medicare and Medicaid Services. 2021. Hospitals data archive 2014-2021. https://data.cms.gov/provider-data/archived-data/hospitals

13. Agency for Healthcare Research and Quality. 2016. Applying the AHRQ Quality Indicators to Hospital Data. https://www.ahrq.gov/sites/default/files/wysiwyg/professionals/systems/hospital/qitoolkit/combined/b1_combo_applyingqis.pdf

14. Rivard, P.E., Elwy, A.R., Loveland, S., Zhao, S., Tsilimingras, D., Elixhauser, A., Romano, P.S. and Rosen, A.K., 2005. Applying patient safety indicators (PSIs) across health care systems: achieving data comparability.

15. Centers for Medicare and Medicaid Services. 2011. LDS Inpatient Data Dictionary. https://www.cms.gov/Research-Statistics-Data-and-Systems/Files-forOrder/LimitedDataSets/Downloads/InpatientVersionJ2011.pdf

16. Peterson, M.D. and Krishnan, C., 2015. Growth charts for muscular strength capacity with quantile regression. American journal of preventive medicine, 49(6), pp. 935–938.

17. Liu, F.T., Ting, K.M. and Zhou, Z.H., 2008, December. Isolation forest. In 2008 eighth ieee international conference on data mining (pp. 413-422). IEEE.

18. Abdullah, S.S., Rostamzadeh, N., Sedig, K., Garg, A.X. and McArthur, E., 2020, June. Visual Analytics for Dimension Reduction and Cluster Analysis of High Dimensional Electronic Health Records. In Informatics (Vol. 7, No. 2, p. 17). Multidisciplinary Digital Publishing Institute.

19. Yan, J., Linn, K.A., Powers, B.W., Zhu, J., Jain, S.H., Kowalski, J.L. and Navathe, A.S., 2019. Applying machine learning algorithms to segment high-cost patient populations. Journal of general internal medicine, 34(2), pp. 211–217.

20. Pearson, K. (1901). “On Lines and Planes of Closest Fit to Systems of Points in Space”. Philosophical Magazine. 2 (11): 559–572. doi:10.1080/14786440109462720.

21. Song, M., Yang, H., Siadat, S.H. and Pechenizkiy, M., 2013. A comparative study of dimensionality reduction techniques to enhance trace clustering performances. Expert Systems with Applications, 40(9), pp. 3722–3737.

22. Ester, M., Kriegel, H.P., Sander, J. and Xu, X., 1996, August. A density-based algorithm for discovering clusters in large spatial databases with noise. In Kdd (Vol. 96, No. 34, pp. 226–231).

23. Schubert, E., Sander, J., Ester, M., Kriegel, H.P. and Xu, X., 2017. DBSCAN revisited, revisited: why and how you should (still) use DBSCAN. ACM Transactions on Database Systems (TODS), 42(3), pp. 1–21.

24. Sander, J., Ester, M., Kriegel, H.P. and Xu, X., 1998. Density-based clustering in spatial databases: The algorithm gdbscan and its applications. Data mining and knowledge discovery, 2(2), pp. 169–194.

25. Van der Maaten, L. and Hinton, G., 2008. Visualizing data using t-SNE. Journal of machine learning research, 9(11).

26. Cieslak, M.C., Castelfranco, A.M., Roncalli, V., Lenz, P.H. and Hartline, D.K., 2020. t-Distributed Stochastic Neighbor Embedding (t-SNE): A tool for eco-physiological transcriptomic analysis. Marine genomics, 51, p. 100723.

27. DeLise, T., 2020. Data Segmentation via t-SNE, DBSCAN, and Random Forest. arXiv preprint 2010.13682.

28. Fujihara, N., Lark, M.E., Fujihara, Y. and Chung, K.C., 2017. The effect of economic downturn on the volume of surgical procedures: a systematic review. International Journal of Surgery, 44, pp. 56’63.

29. Tonna, J.E., Hanson, H.A., Cohan, J.N., McCrum, M.L., Horns, J.J., Brooke, B.S., Das, R., Kelly, B.C., Campbell, A.J. and Hotaling, J., 2020. Balancing revenue generation with capacity generation: case distribution, financial impact and hospital capacity changes from cancelling or resuming elective surgeries in the US during COVID-19. BMC health services research, 20(1), pp. 1–7.

